# Remote fingerstick blood collection for SARS-CoV-2 antibody testing

**DOI:** 10.1101/2020.10.27.20221028

**Authors:** Wilfredo F. Garcia-Beltran, Tyler E. Miller, Grace Kirkpatrick, Andrea Nixon, Michael G. Astudillo, Diane Yang, Lisa M. Mahanta, Mandakolathur Murali, Anand Dighe, Jochen Lennerz, Julia Thierauf, Vivek Naranbhai, A. John Iafrate

**Affiliations:** Department of Pathology, Massachusetts General Hospital, Boston, MA; Department of Medicine, Massachusetts General, Hospital, Boston, MA; Mass General Brigham Biobank, Boston, MA

## Abstract

The rapid worldwide spread of SARS-CoV-2 infection has propelled the accelerated development of serological tests that can detect anti-SARS-CoV-2 antibodies. These have been used for studying the prevalence and spread of infection in different populations, helping establish a recent diagnosis of COVID-19, and will likely be used to confirm humoral immunity after infection or vaccination. However, nearly all lab-based high-throughput SARS-CoV-2 serological assays require a serum sample from venous blood draw, limiting their applications and scalability. Here, we present a method that enables large scale SARS-CoV-2 serological studies by combining self or office collection of fingerprick blood with a volumetric absorptive microsampling device (Mitra, Neoteryx, LLC) with a high-throughput electrochemiluminescence-based SARS-CoV-2 total antibody assay (Roche Elecsys, Roche Diagnostics, Inc.) that is EUA approved for use on serum samples and widely used by clinical laboratories around the world. We found that the Roche Elecsys assay has a high dynamic range that allows for accurate detection of SARS-CoV-2 antibodies in serum samples diluted 1:20 as well as contrived dried blood extracts. Extracts of dried blood from Mitra devices acquired in a community seroprevalence study showed near identical sensitivity and specificity in detection of SARS-CoV-2 antibodies as compared to neat sera using predefined thresholds for each specimen type. Overall, this study affirms the use of Mitra dried blood collection device with the Roche Elecsys SARS-CoV-2 total antibody assay for remote or at-home testing as well as large-scale community seroprevalence studies.

## INTRODUCTION

The ongoing pandemic of coronavirus disease 2019 (COVID-19), caused by infection with severe acute respiratory system coronavirus 2 (SARS-CoV-2), has continued to ignite the rapid development of diagnostic tests that can detect active or past infection. To date, molecular tests that detect viral RNA are the gold standard for diagnosing and reporting active infection. However, detection of antibodies to SARS-CoV-2 via serological assays is necessary to identify recently infected individuals. This detection is critical for studies that measure the prevalence and spread of infection and is likely to be used to confirm the development of humoral immunity after infection or vaccination. Currently, there are over 30 emergency use authorization (EUA)-approved serological tests and there currently exists no wide-spread or gold-standard platform^1^. Several studies have compared the performance of these various platforms, including: Euroimmun anti-spike IgG enzyme-linked immunosorbent assay (ELISA), Bio-Rad Platelia anti-nucleocapsid total antibodies ELISA, Wantai anti-spike total antibodies ELISA, Diasorin Liaison anti-spike IgG chemiluminescence immunoassay (CLIA), Abbott Alinity anti-nucleocapsid IgG chemiluminescent microparticle-based immunoassay (CMIA), Siemens anti-spike IgG CMIA, and Roche Elecsys anti-nucleocapsid total antibodies electrochemiluminescence-based immunoassay (ECLIA)^2–4^. Based on our previous work, we chose at our institution to deploy the Roche Elecsys SARS-CoV-2 total antibody ECLIA that detects IgG, IgM, and/or IgA antibodies to SARS-CoV-2 nucleocapsid^2^, which many other centers around the world have also done^5^. While this has provided great benefit to diagnosing recent SARS-CoV-2 infection for inpatients and outpatients, this test requires collection of serum from whole blood via phlebotomy, as do the other EUA-approved serological tests mentioned above. This exposes clinical staff to potentially infectious patients and limits the scalability of SARS-CoV-2 antibody testing due to phlebotomy requirements.

Consequently, many investigators have searched for a strategy that involves remote collection or self-collection of serum. Several methods of remote blood collection have long existed and have been used to detect SARS-CoV-2 antibodies, including dried blood spot (DBS) cards^6,7^, yet extraction of serum from these specimens is cumbersome and low throughput. To address these shortcomings and enable high-throughput processing, Neoteryx, LLC developed a novel device called Mitra. This device uses an absorptive tip that collects a standardized volume of fingerprick blood and can be stored long term under dry conditions. This allows for remote self-collected blood that can be mailed back to a central facility and processed through a high-throughput extraction pipeline in 96-well plate format. Accordingly, the National Institute of Health (NIH) has partnered with the developers of this device and have validated its use with an in-house-developed serological assay that measures IgG and IgM antibodies to SARS-CoV-2 spike protein^8^, and have an ongoing clinical trial assessing the sero-prevalence of SARS-CoV-2 infection in different areas of the country^9^.

In this study, we assess the performance of the Mitra device pipelined into the Roche Elecsys SARS-CoV-2 antibody assay to expand our SARS-CoV-2 testing capability to at-home testing and large-scale community seroprevalence studies.

## METHODS

### Dried blood collection of contrived blood, whole blood, and fingerprick blood samples

Contrived whole blood was generated by mixing serum with an equal volume of type O-negative packed red blood cells (hematocrit of approximately 80%). Whole blood was obtained from discarded segments (tubing) from whole blood donations. Fingerprick blood was expressed using a spring-loaded lancet (Becton Dickinson). 20-μL Mitra devices (Neoteryx, LLC) were used to collect fingerprick and blood sample specimens, followed by dry storage for at least 24 h. Mitra is a Food and Drug Administration (FDA) class I exempt medical device registered with the FDA.

### Dried blood extraction

Mitra devices containing dried blood samples were placed in a deep (2-mL) 96-well plate with a fitted holder (Neoteryx, LLC) that allows for the devices to be slightly suspended. The deep 96-well plates contained 200 μL/well of extraction buffer, which consisted of 1% BSA, 50 mM Tris (pH 8.0), 140 mM NaCl, and 0.05% Tween-20. Our extraction buffer contained BSA to help stabilize antibodies as well as 0.02% Tween-20 to help facilitate extraction and inactivate any potentially infectious virus^10^. Plates were then placed on a shaker for 2 h at 600 RPM. Devices were subsequently removed, leaving 200 μL of dried blood extract in each well. Of note, extracts exhibited significant hemolysis (dark red in color).

### Roche Elecsys SARS-CoV-2 total antibody assay

We used the electrochemiluminescence-based Roche Elecsys anti-SARS-CoV-2 immunoassay, which detects IgG, IgM, and/or IgA antibodies to the SARS-CoV-2 nucleocapsid protein. The assay was run on a Roche Cobas 8000 e801 Immunoassay Analyzer, which also measured hemolysis and lipemia indices. A minimum volume of approximately 200 μL of sample per run was required when using a tube insert.

### Patient samples

Use of patient samples for the development and validation of SARS-CoV-2 diagnostic tests was approved by Partners Institutional Review Board (protocol 2020P000895).

## RESULTS

In order to determine the feasibility of running dried blood extracts from 20-μL Mitra devices on the Roche Elecsys SARS-CoV-2 total antibody assay, we first tested serum samples from COVID-19 patients (confirmed by nasopharyngeal swab polymerase chain reaction) diluted 1:20 in extraction buffer. A dilution of 1:20 was chosen based on a conservative estimate that assumes that of the 20 μL of whole blood that dries onto 20-μL Mitra device, there will be at least 10 μL of dried serum extracted into 200 μL of extraction buffer. Analysis of neat serum versus 1:20-diluted serum from 72 COVID-19 patients showed a positive, non-linear correlation of cut-off index (COI) values measured by the Roche Cobas 8000 e801 Immunoassay Analyzer as well as a high dynamic range of detection with COI ranging from approximately 0.08 to 140 (**Figure 1A**). Using the threshold of 1.0 established by the manufacturer, neat serum samples showed 86% (62/72) seropositivity while diluted serum samples showed 46% (33/72) seropositivity. However, adjusting the threshold to 0.15, the seropositivity of diluted serum samples increased to 88% (63/72). Previous studies have demonstrated that this adjustment in the COI threshold for the Roche assay can increase the sensitivity of SARS-CoV-2 antibody detection without sacrificing specificity^11^.

**Figure 1:**
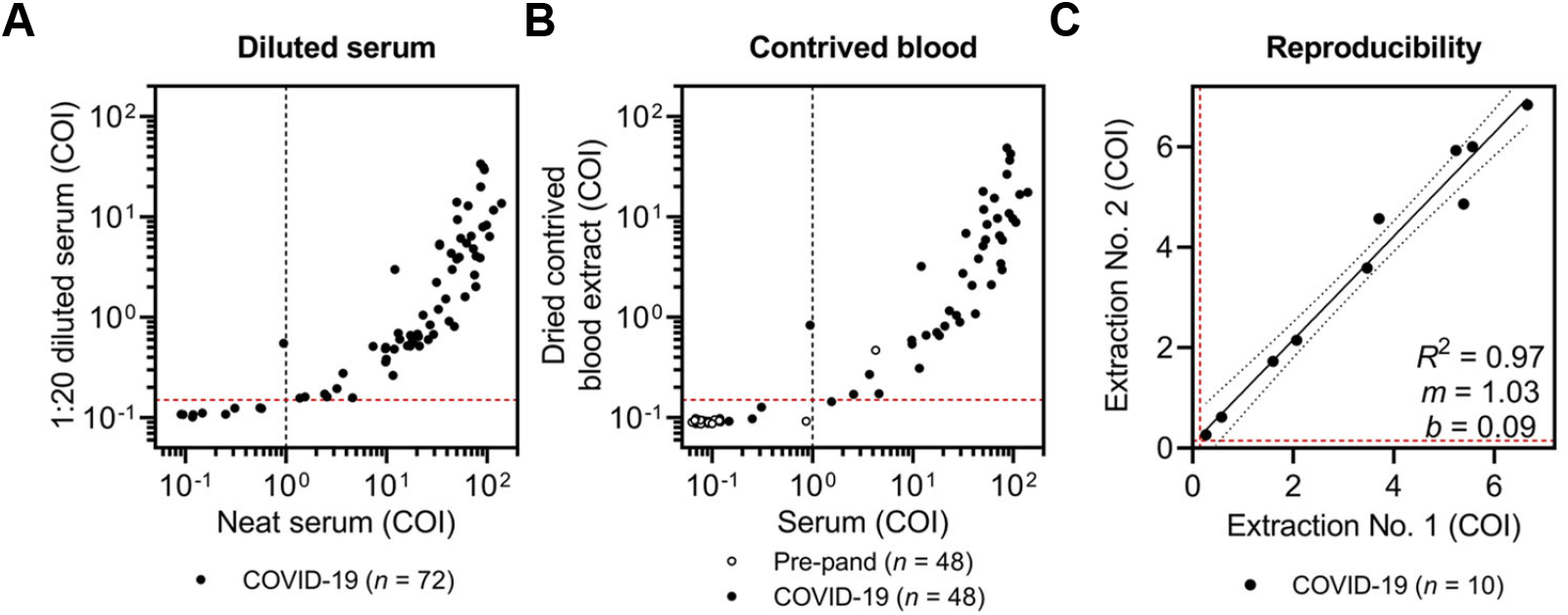
Roche Elecsys SARS-CoV-2 total antibody assay can accurately detect antibodies in diluted serum samples and dried blood extracted from Mitra devices. (**A**) Serum samples from COVID-19 patients (*n* = 72) were run as neat serum as well as serum diluted 1:20 in extraction buffer on the Roche Elecsys SARS-CoV-2 total antibody assay on the Roche Cobas 8000 instrument, with cut-off index (COI) values reported. (**B**) Contrived blood from COVID-19 patients (*n* = 48) and pre-pandemic individuals (*n* = 48) was collected and stored on Mitra devices and subsequently extracted and run on the Roche Elecsys assay along with paired neat serum. (**C**) Duplicate Mitra devices were used to collect contrived blood from COVID-19 patients (*n* = 10), and extractions were performed in two separate facilities and run on the same Roche Elecsys assay and Roche Cobas 8000 instrument; Pearson correlation resulted in the following: coefficient of determination (*R*^2^) = 0.97, slope (*m*) = 1.03, and intercept (*b*) = 0.09. For all, the dotted black line indicates the manufacturer-established COI threshold of 1.0 (for serum) and the dotted red line indicates the new COI threshold of 0.15 (for diluted or extracted samples).

Having demonstrated that the Mitra-Roche assay can accurately detect SARS-CoV-2 antibodies in diluted serum samples, we proceeded to generate extracts from dried blood collected on Mitra devices using contrived whole blood with known serostatus. For this, we mixed serum samples from 48 COVID-19 patients and 48 pre-pandemic individuals with an equal volume of O-negative donor packed red blood cells to mimic whole blood, which was subsequently collected onto 20-μL Mitra devices. After 24 hours of dry storage, samples were extracted and run on the Roche Elecsys assay. Quality control measurements made by the Roche Cobas instrument of hemolysis and lipemia were included, and each extracted sample, which was dark red in color, unsurprisingly contained high hemolysis and lipemia indices due to the nature of the sample (data not shown). Nevertheless, this did not affect the SARS-CoV-2 antibody assay results, and using a threshold of 1.0 for neat serum and 0.15 for extracted samples, a concordance of 98% (94/96) was achieved, with the only two discordant specimens being COVID-19 patient samples (**Figure 1B**). Of the two discrepant COVID-19 patient samples, one was positive by the neat serum specimen, but just below the threshold by the dried blood specimen, and the other was positive by the dried blood specimen but just below the threshold by the neat serum specimen. In addition, a false positive pre-pandemic specimen, which was selected as one of two pre-pandemic specimens that were false positive in the Roche assay in a cohort of 1,200 pre-pandemic specimens, was still detected in the extract sample, indicating that false positive signals indeed dilute with sample dilution, and could reflect truly cross-reactive antibodies. We also generated duplicate Mitra collections devices from contrived whole blood and performed extractions in two independent facilities and found high precision (**Figure 1C**, *R*^2^ = 0.97), highlighting the reproducibility of this testing strategy.

To test the performance of the combined Mitra-Roche assay on low- and high-seroprevalence cohorts during the pandemic, we collected specimens from pre-screened blood donors at the Massachusetts General Hospital Blood Donor Center as well as in a community seroprevalence study conducted in Chelsea, Massachusetts, the densest and most affected area for SARS-CoV-2 infection^12^. For blood donors, we collected serum and dried venous blood on Mitra devices and found that the assay showed a seroprevalence of 1% (1/143) with 100% concordance between serum and extracted samples (**Figure 2A**). For community samples, we collected dried fingerprick blood on Mitra devices and serum from antecubital vein phlebotomy in 402 participants. Dried fingerprick blood extracts and sera were run on the Roche Cobas instrument, which revealed a seroprevalence of 24% (98/402) in serum samples, 24% (97/402) in extracted sample, and a concordance of 99% (397/402) (**Figure 2B**). Interestingly, we saw a threshold effect in neat serum samples, with extracted samples that showed the highest COI demonstrating slightly decreased COI in the neat serum, most likely representing a ‘hook effect’ (**Figure 2C**). This effect, however, did not decrease COI sufficiently to approximate the COI threshold, and thus is unlikely to affect Roche results as a qualitative read-out.

**Figure 2:**
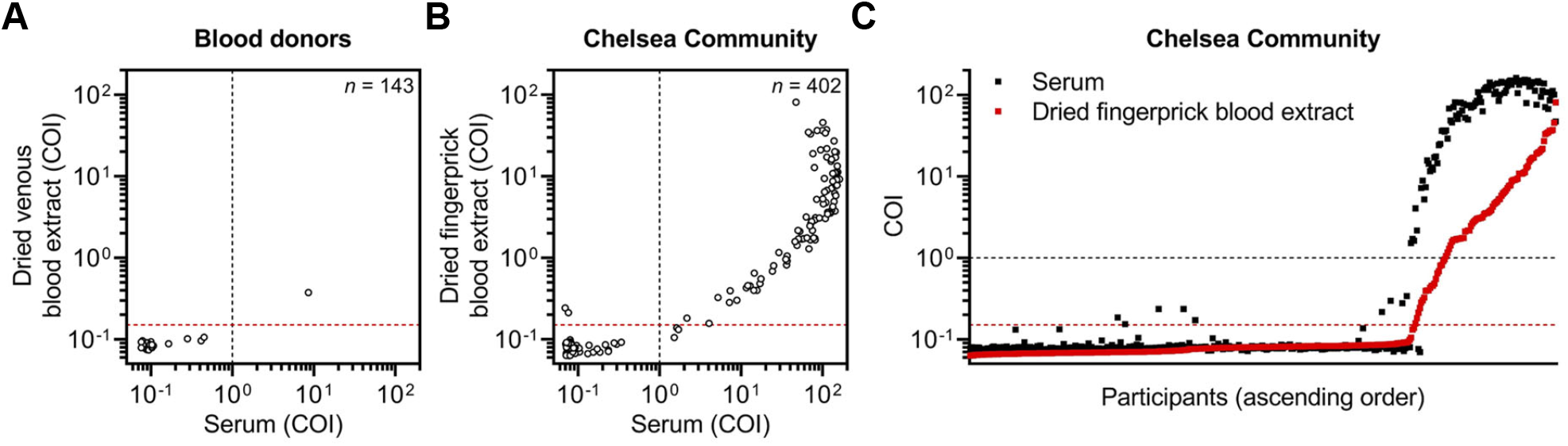
Mitra-Roche assay serves as a high-throughput platform for seroprevalence studies. (**A**) Serum and dried venous blood extracted from Mitra devices were collected from pre-screened blood donors (*n* = 143) and run on the Roche Elecsys assay. (**B**) Serum from antecubital vein phlebotomy and dried fingerprick blood extracted from Mitra devices were collected from participants in a Chelsea community seroprevalence study (*n* = 407) an run on the Roche Elecsys assay. (**C**) COI values from the serum (black squares) and dried fingerprick blood extracted from Mitra devices (red squares) for each participant are overlaid and presented in ascending order (according to COI from dried fingerprick blood extract). For all, the dotted black line indicates the manufacturer-established COI threshold of 1.0 for serum and the dotted red line indicates the new COI threshold of 0.15 for extracted samples.

## DISCUSSION

The need for remote and large-scale testing for SARS-CoV-2 antibodies is increasing as the population of convalescent individuals continues to increase and vaccines are soon to be implemented. In this study, we validate the use of a volumetric absorptive microsampling device (Mitra) for fingerprick blood on a widely used, EUA-approved SARS-CoV-2 antibody test (Roche Elecsys). Currently, the Roche Elecsys test detects IgG, IgM, and/or IgA antibodies to SARS-CoV-2 nucleocapsid protein, an antigen that is not present in most SARS-CoV-2 vaccines. However, a Roche Elecsys assay that detects antibodies to SARS-CoV-2 spike is currently being developed^13^ and will be useful to detect immune responses to vaccines, all of which contain all or a portion of the spike antigen. Both tests together can even help discriminate individuals who were infected with circulating virus (i.e. positive for both nucleocapsid and spike antibodies) or administered one of the vaccines being developed (i.e. negative for nucleocapsid antibodies but positive for spike antibodies). As such, this protocol and collection method, which is readily adaptable to the upcoming version of Roche Elecsys assays, can enable remote confirmation of vaccination responses. Currently, the NIH is using this collection method in conjunction with an in-house developed serological assay, but our study validates its use in facilities already deploying the Roche Elecsys SARS-CoV-2 total antibody assay. In addition, although performance may not be guaranteed due to potential interferences from extracted dried blood, this study provides a straightforward framework for validating other SARS-CoV-2 serological testing platforms, such as from Euroimmun, Bio-Rad, Diasorin, Abbott, and Siemens. Overall, we believe these findings to be of significance for clinical laboratories around the world as we combat this pandemic.

## Data Availability

De-identified data can be made available upon request.

## ACKNOWLEDGEMENTS

We wish to thank Dr. Robert S. Makar for providing whole blood samples from blood donors. We also thank Dr. Edward T. Ryan for providing pre-pandemic serum samples.

## AUTHOR CONTRIBUTIONS

W.F.G.B., T.E.M., M.G.A., A.J.I., L.M.M., and A.D. designed the experiments. W.F.G.B., T.E.M., G.K., A.N., M.G.A., D.Y., and L.M.M. carried out experiments and analyzed data. J.T. and V.N. helped design and carry out experiments and studies and provided useful discussion. M.M. and J.L. provided key insights and useful discussions into experimental and study design. W.F.G.B. and T.E.M. wrote the manuscript with contributions from all authors.

## REFERENCES

1. Center for Devices, Radiological Health. EUA Authorized Serology Test Performance. Published 2020. Accessed October 17, 2020. https://www.fda.gov/medical-devices/coronavirus-disease-2019-covid-19-emergency-use-authorizations-medical-devices/eua-authorized-serology-test-performance

2. Turbett SE, Anahtar M, Dighe AS, et al. Evaluation of Three Commercial SARS-CoV-2 Serologic Assays and their Performance in Two-Test Algorithms. J Clin Microbiol. [Published online ahead of print October 5, 2020]. doi:10.1128/JCM.01892-20

3. Brochot E, Demey B, Handala L, François C, Duverlie G, Castelain S. Comparison of different serological assays for SARS-CoV-2 in real life. J Clin Virol. 2020;130:104569.

4. National SARS-CoV-2 Serology Assay Evaluation Group. Performance characteristics of five immunoassays for SARS-CoV-2: a head-to-head benchmark comparison. Lancet Infect Dis. Published online September 23, 2020. doi:10.1016/S1473-3099(20)30634-4

5. Roche to launch laboratory SARS-CoV-2 antigen test to support high-volume testing of suspected COVID-19 patients. Accessed October 20, 2020. https://www.roche.com/media/releases/med-cor-2020-10-13.htm

6. Morley GL, Taylor S, Jossi S, et al. Sensitive Detection of SARS-CoV-2-Specific Antibodies in Dried Blood Spot Samples. Emerg Infect Dis. 2020;26(12). doi:10.3201/eid2612.203309

7. McDade TW, McNally EM, Zelikovich AS, et al. High seroprevalence for SARS-CoV-2 among household members of essential workers detected using a dried blood spot assay. PLoS One. 2020;15(8):e0237833.

8. Klumpp-Thomas C, Kalish H, Drew M, et al. Standardization of enzyme-linked immunosorbent assays for serosurveys of the SARS-CoV-2 pandemic using clinical and at-home blood sampling. medRxiv. [Preprint - Published online May 25, 2020.] doi:10.1101/2020.05.21.20109280

9. NIH Begins Study to Quantify Undetected Cases of Coronavirus Infection. Accessed October 17, 2020. http://www.niaid.nih.gov/news-events/nih-begins-study-quantify-undetected-cases-coronavirus-infection

10. Mayo DR, Beckwith WH 3rd. Inactivation of West Nile virus during serologic testing and transport. J Clin Microbiol. 2002;40(8):3044–3046.

11. Favresse J, Eucher C, Elsen M, Tré-Hardy M, Dogné J-M, Douxfils J. Clinical Performance of the Elecsys Electrochemiluminescent Immunoassay for the Detection of SARS-CoV-2 Total Antibodies. Clin Chem. 2020;66(8):1104–1106.

12. Naranbhai V, Chang CC, Garcia-Beltran WF, et al. High seroprevalence of anti-SARS-CoV-2 antibodies in Chelsea, Massachusetts. J Infect Dis. Published online September 9, 2020. doi:10.1093/infdis/jiaa579

13. Elecsys Anti-SARS-CoV-2 S. Accessed October 21, 2020. https://diagnostics.roche.com/global/en/products/params/elecsys-anti-sars-cov-2-s.html

